# Dietary Patterns Among 13-17-Years Old School Adolescents: A National Comparison of the Three Geographical Regions in Bolivia

**DOI:** 10.1101/2021.07.22.21259344

**Authors:** Noelia Villalta, Nattalia Arauco, Marisol Mamani, Daniel E Illanes, Pramil Singh, Lenildo De Moura, Joan Sabaté

## Abstract

**Introduction:** Bolivia is considered one to the most biodiverse countries in the world. It has three ecological floors or geographical regions (highlands, plains and valleys) with very different climatic and cultural characteristics. Traditionally dietary patterns varied in each region. Because two-thirds of premature deaths are associated with behaviors that begin in adolescence, understanding dietary patterns among adolescents is important to improving health. The aim of this study was to identify dietary patterns of adolescents in the different geographical regions of Bolivia.

**Methodology:** Data were obtained from the World Health Organization’s Global School-based Student Health Survey 2018 that assesses risky behaviors and protective factors in students aged 13 to 17 years old. It uses an observational cross-sectional method and included 7945 adolescents in schools that were representative of all students in this age group in each of the three geographical regions in Bolivia.

**Results:** 3.9% of students in Bolivia are food insecure, with the highest percentage in the highlands (4.6%) and the lowest in the plains (3.1%). Adolescents in the highlands consume vegetables and fruits more frequently than adolescents in the plains and valleys. Water consumption in the valleys was 19.6% students who drink 1-3 glasses/d. Dairy consumption ≥2 in 7 days is higher in the highlands and lower in the valleys. Consumption of soft drinks at ≥1 glass per day is 34.3% in the plains and 30.1% in the valleys. Junk food consumption ≥1 in 7 days was 62.0% with a greater presence on the plains at 65.7%.

**Conclusion:** The highlands revealed a greater presence of protective factors than the other regions. The valley, despite having greater access to fruits and vegetables, has a greater presence of risky behaviors. Teenagers in the plains have greater exposure to advertising and access to junk food at school.

## INTRODUCTION

Most countries in Latin America are undergoing socioeconomic changes, accelerated urbanization, greater availability and access to processed foods, coupled with poor health education. These factors have been proved to have a great influence on eating patterns and health behaviors, leading to low consumption of vegetables and fruits and high consumption of highly processed foods (1–3). Latin America has the highest prevalence of obesity in the world and it is increasing rapidly (4,5). The complex interaction of the associated factors that led to this situation highlights the need to make greater efforts to understand them in order to propose evidence-based interventions. The current data indicate that adolescents who are overweight due to inadequate lifestyles may maintain these habits through adulthood, putting them at risk of developing a wide range of diseases especially chronic diseases (6). Schools play an important part in the development of healthy eating habits in adolescents. Studies have shown that the more proximity to junk food in the school stores, the more junk food is purchased, leading to poorer nutritional status in adolescents (6,7).

The Global School Health Survey was designed by the World Health Organization to obtain systematic information from students to provide accurate data on health behaviors and protective factors among students (8). The Global School Health Survey (GSHS) is an important part of the World Health Organization’s surveillance system to prevent and control non-communicable diseases and their risk factors. A total of 33 countries in the Region of the Americas have applied this survey (9). The survey was first conducted in Bolivia in 2012 by the Ministry of Health and the Pan American Health Organization (10). A second survey has conducted in 2018. Based on data from the latter, the aim of this article is to describe the eating patterns and consumption determinants of selected foods and drinks of schooled adolescents in Bolivia and its three geographically and culturally diverse regions: the highlands, the valleys and the plains.

## METHODS

The Global School Health Survey (GSHS) is a cross-sectional study conducted in schools among students from 13 to 17 years old. The questions were chosen, translated, adapted, and culturally validated, respecting the colloquial language of the region. The instrument is an anonymous, self-administered questionnaire with core modules, core-expanded questions, and country-specific questions. Since this is a self-reported instrument information may be susceptible to recall bias. The project was approved by the Ministry of Health Review Board and the Health Investigation Institute of Bolivia. Parental consent was obtained via letters sent to students homes from schools, with the cooperation of the Ministry of Education.

### Sample

The sample was taken from the nine departments in Bolivia, which are divided into three geographical regions (known locally as ecological floors): the highlands (La Paz, Oruro, Potosí), the valleys (Cochabamba, Chuquisaca, Tarija), and the plains (Pando, Beni, Santa Cruz). Each of these regions varies from a height of 3 600 - 4 100 meters above the sea in the highlands, 1 800 – 3 000 m.a.s.l. in the valleys, and 200 – 400 m.a.s.l. in the plains. These characteristics define the ecosystem and climate that results in different patterns in farming and foods that adolescents can access in each region (11,12). Furthermore, the sociocultural, socio-demographic, in the three regions are very different.

The GSHS uses a standardized scientific sample selection process. It uses clusters of students calculated by probability proportional to size method and systematic sampling (13). Schools were selected through probability proportional to enrollment size, while classes in each selected school were selected through random sampling. Data included 9 467 students from 96 public and private schools from the urban and rural areas, corresponding to 32 schools in each geographical region. It is representative of the nation and of each region, which will allow us to describe group and national patterns of eating behaviors in adolescents.

### Data Collection

Data collection was coordinated by the Ministry of Health through the MI SALUD program, the National Institute of Health Research (IINSAD), and the Pan American Health Organization; The Survey, conducted at schools, used a self-administered questionnaire that was administered during one regular class period by two qualified persons in each class. Among the 10 modules of the questionnaire, 18 questions related to dietary patterns such as “During the past 7 days, how many times did you drink a can, bottle, or glass of a sugar-sweetened beverage?” The complete module is available as Appendix 1.

Coordinators, health professionals and university students were trained to implement the survey. This training included two workshops that are provided to Survey Coordinators from the participating country; the first workshop builds the capacity of Survey Coordinators to implement the survey, the second workshop covers the administration procedures that ensure that the survey data are of the highest quality. In this stage, health students and MI SALUD professionals learned the correct way to interpret each question and to solve any problem the students may have. The survey was administered between August 13 and 24 of 2018.

## STATISTICAL ANALYSIS

Since GSHS surveys employ a two-stage sampling design, to analyse GSHS data correctly, three variables are required to be used: weight, stratum, and PSU for every student record in the GSHS. The weights allow the results to be generalized to the study population and the national student population. The stratum reflects the first stage of the GSHS sampling (school level), and the PSU reflects the second stage (classroom level) assigned sequentially starting, with the class rooms in the schools with the largest enrolment of students and continuing through the list to the class rooms in the schools with the smallest enrolment. GSHS methodology recommends that statistical software packages that account for this sampling design must be used to ensure the quality of the analysis. This study used Epiinfo to report weighted prevalence with 95% Confidence Interval. Missing data was not statistically imputed.

## RESULTS

### Response

A total 7 931 students participated in the Bolivia GSHS; the student response rate was 84%, with variability in the response in each in the geographical regions. The highest response was in the highlands, and lowest in the valleys (Table 1) where some students chose not to participate.

**Table 1.**
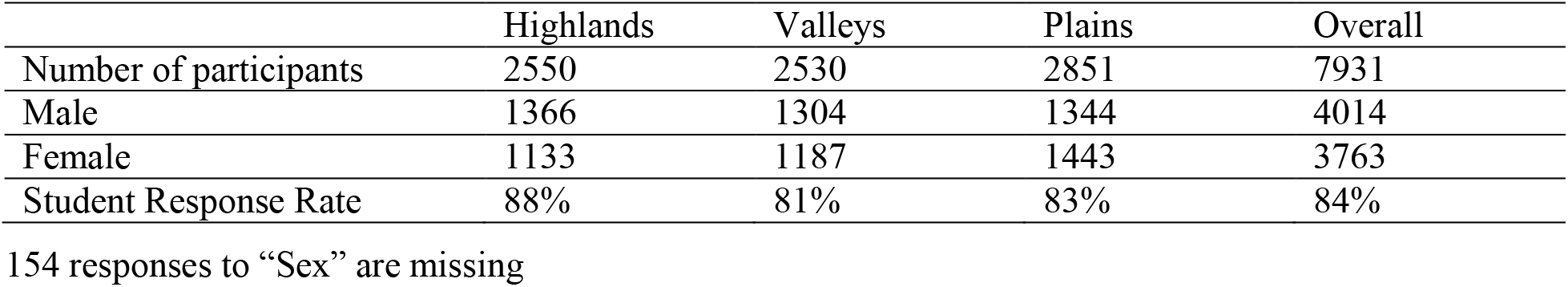
Study sample of 13-17 year old Bolivian student responses to Global School-based Student Health Survey (GSHS) in 2018

### Bolivia school-going adolescents eating habits pattern

#### NCD protective factors

This group includes: Students who ate breakfast most of the time or always (53,2%), students who ate vegetables three or more times per day (21,6%), students who ate fruits two or more times per day (26,2%), students who reported water as their favorite drink when they are thristy (62,1%), and water consumption two or three times per day (21,0%) (Figure 1-A). The regional comparative figure show a similar pattern in the different variables. However, there are slightly more protective factors in the highlands, except in the preference of water as a favorite drink when thirsty (Figure 1-B).

**Figure 1.**
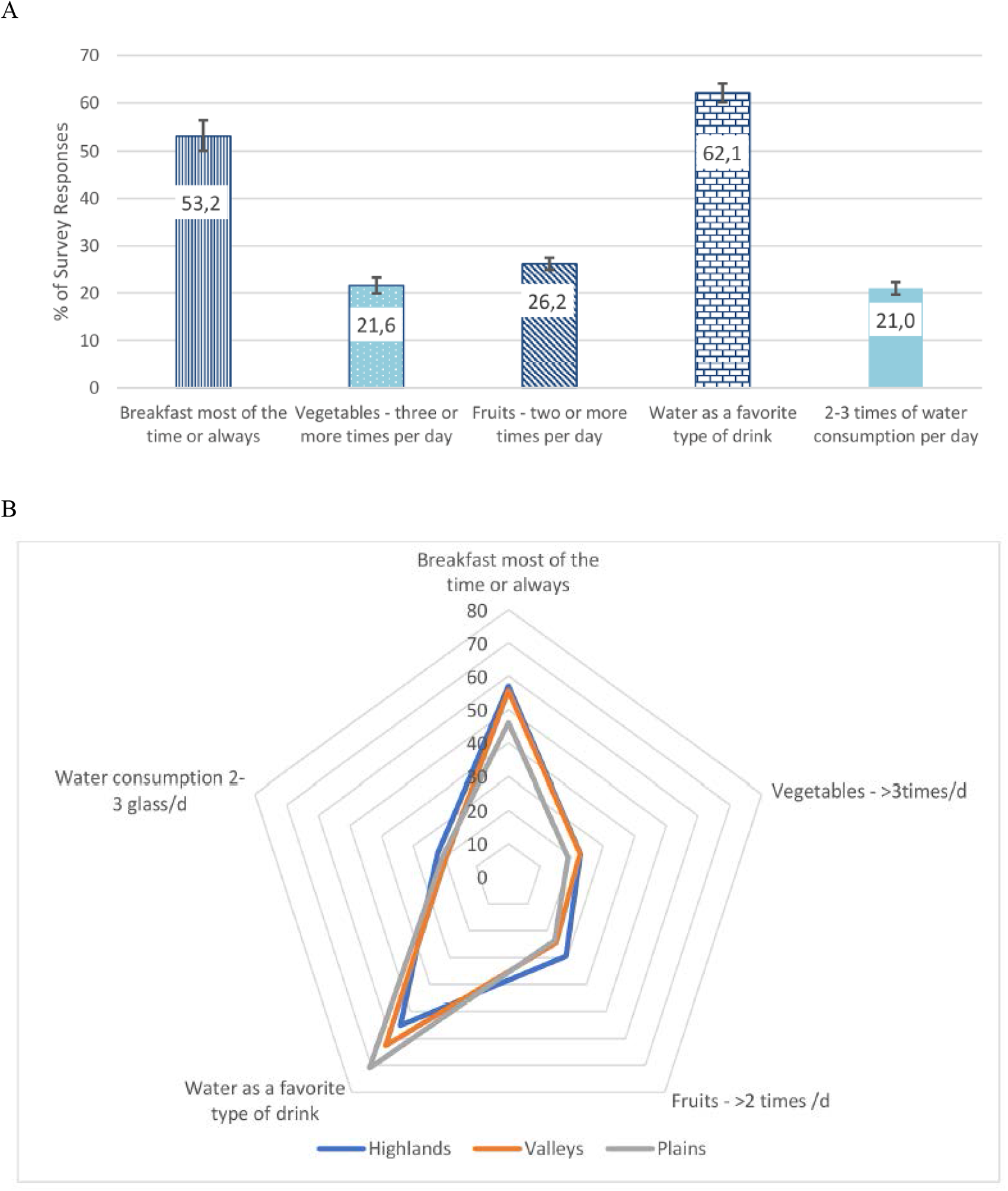
NCD Protective factors (A) and regional patterns (B) in 13 – 17-year-old Bolivian students

#### NCD risk factors

Included in this group are: Students who drank 1 or more carbonated drinks per day during the last seven days (33,1%); students who ate high-fat foods two or more times per day (8,3%); students who can buy or get free carbonated soft drinks in school (34,9); students who can buy or get free junk food in school (37,8%); and students who saw a lot of advertisement for junk food and carbonated drinks in school (14,9%) (Figure 2-A). Regarding the regional comparative figure, a similar pattern is evident for all three regions. Nonetheless, as in the figure above, there is less presence of these factors in the highlands (Figure 2-B).

**Figure 2.**
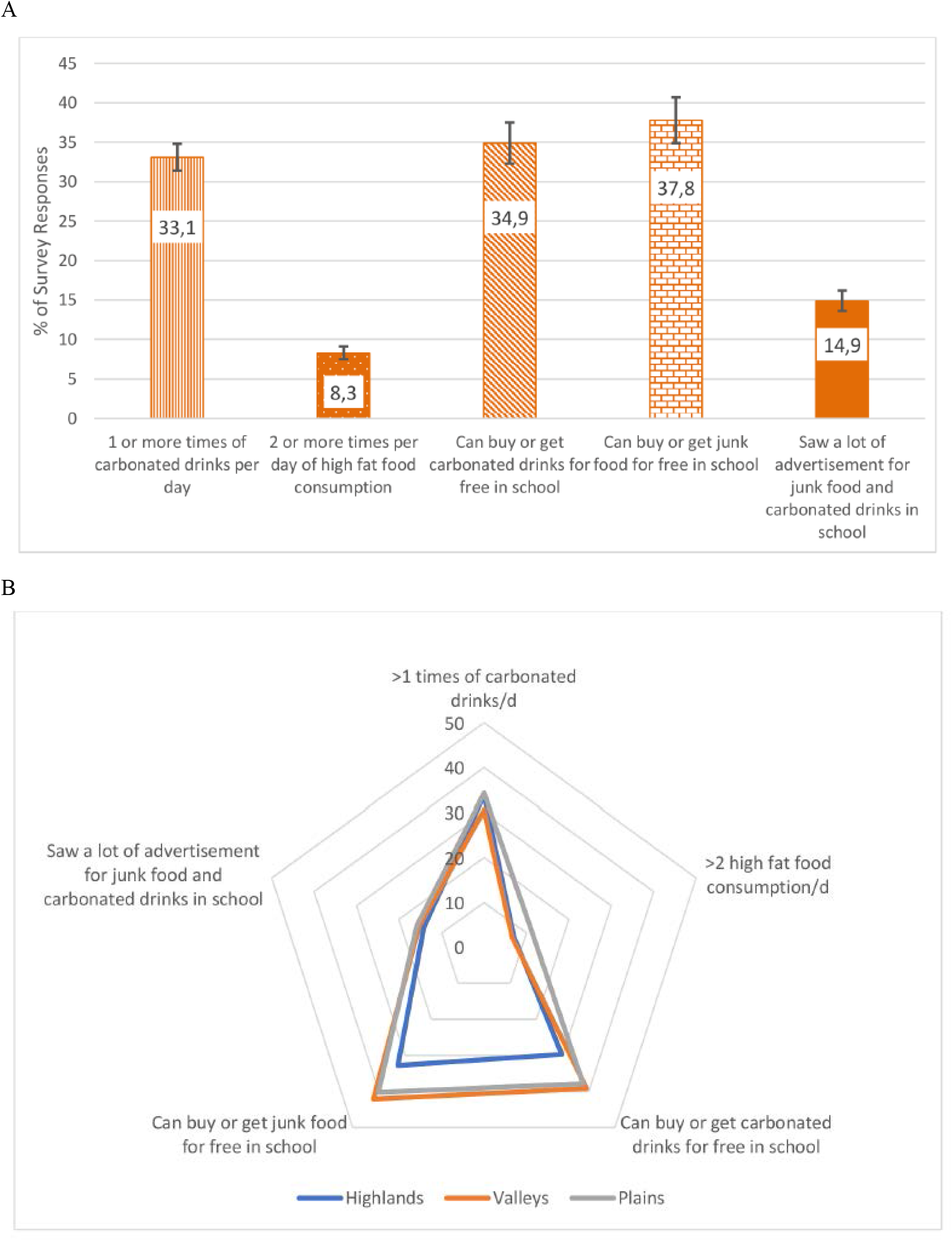
NCD risk factors (A) and regional patterns (B) in 13 – 17-year-old Bolivian students

### Adolescent eating habits regional comparative data

#### Breakfast consumption

The question was directed to the frequency of eating breakfast in the last month. The highest frequency in the highlands is registered in the “ate breakfast always or most of the time category” with 56,4% and only a 6,1% who never ate breakfast in the last month. The valleys registered a similar pattern. In the plains, the highest frequency was for students who ate breakfast sometimes with 46,9% (Figure 3).

**Figure 3.**
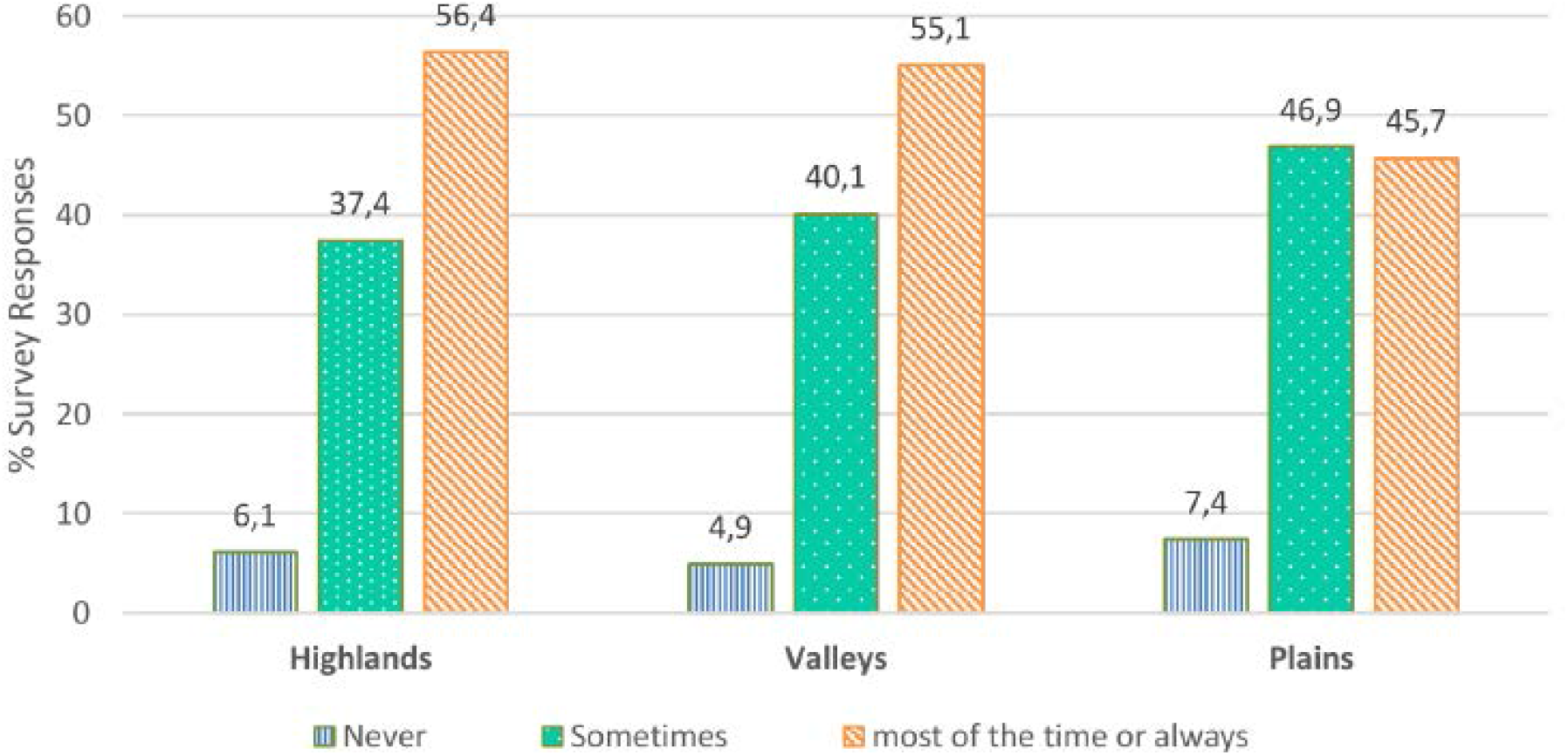
Breakfast consumption patterns in 13 – 17-year-old Bolivian students by geographic region

#### Fruit consumption per week

the three regional patterns have similar results; the highest frequency in daily consumption per week is in the highlands with 44.2%. In contrast, the plains register the highest percentage of adolescents who didn’t eat fruits in the last seven days with 14,1%. With regard to **Vegetable consumption per week**, 18.6% of the adolescents in the highlands consumed vegetables 1-3 times per week while 56.4% ate vegetables daily. The highest percentage of adolescents that did not eat vegetables is in the plains with 6.8% (Figure 4).

**Figure 4.**
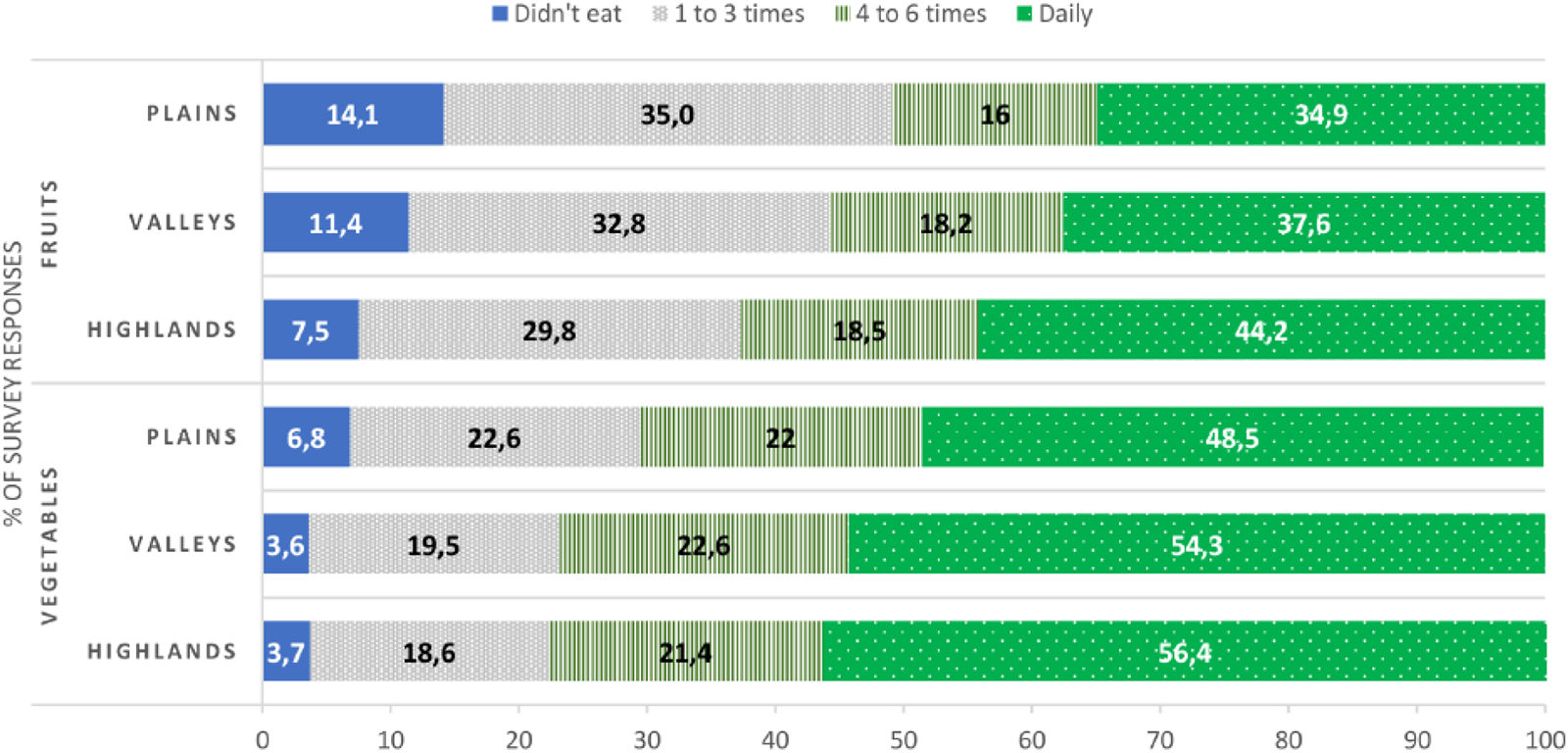
Vegetable and fruit consumption patterns in 13 – 17-year-old Bolivian students by geographic region

#### Favorite type of drink when thirsty

The adolescents in the plains preferred water when thirsty (71,0%), followed by the valleys with 62,2%. Carbonated drinks are more preferred by the adolescents in the highlands with 18,4 %. The category “Refrescos” which is a traditional flavored water (the question included the examples: “Mocochinchi and Somó”) had the highest presence in the highland adolescents (Figure 5-A).

**Figure 5.**
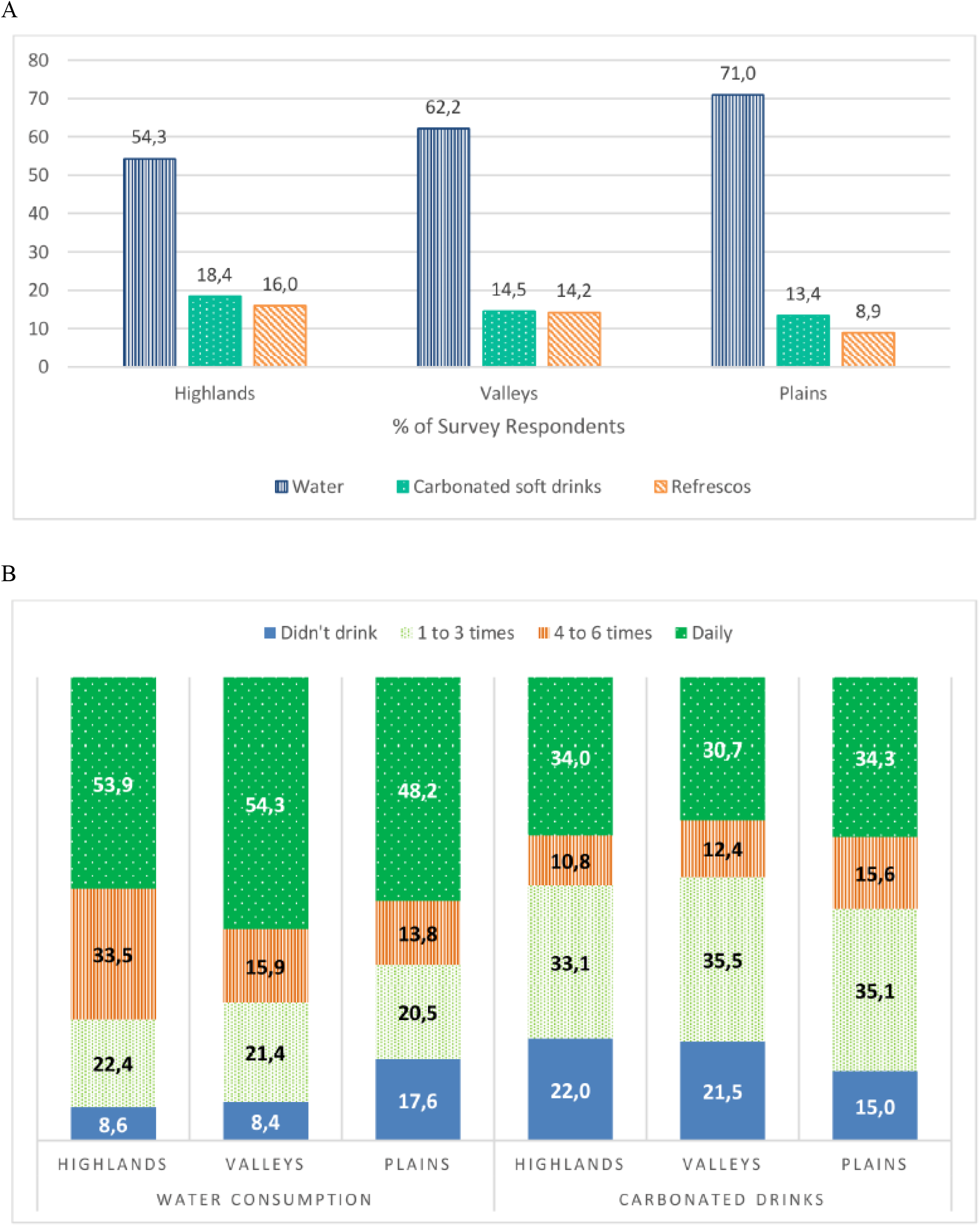
Favorite drink when thirsty (A) and water and carbonated drinks consumption (B) of 13 – 17-year old Bolivian students by geographic region

#### About water and carbonated soft drinks consumption per week

Generally, the students of the three geographical regions consume more carbonated drinks than water. It is important to note that the valleys register the highest daily water consumption whereas the plains have the lowest rate (Figure 5-B).

#### Learning about healthy nutritional habits in school during this school year

The results show that schools in the plains give more importance to teaching about healthy nutritional habits, and the lower frequency in teaching is in the highlands (Table 2).

**Table 2.** Healthy nutrition habits learned in school by 13-17 year geographic region

|  | Highlands |  | Valleys |  | Plains |  |
| --- | --- | --- | --- | --- | --- | --- |
|  | Yes | No | Yes | No | Yes | No |
| Benefits of healthy eating | 78,9 | 11,9 | 81,4 | 11,3 | 84,8 | 8,1 |
| Benefits of eating more fruits and vegetables | 80,5 | 12,0 | 83,4 | 10,4 | 87,1 | 8,3 |
| Safe food preparation and storage |  |  |  |  |  |  |

#### Advertising junk food and carbonated drinks on television, the internet and text messages

The adolescents of the all three regions have seen more junk food and carbonated soft drink advertisements through television and a lower percentage of advertising through text messages (Figure 6).

**Figure 6.**
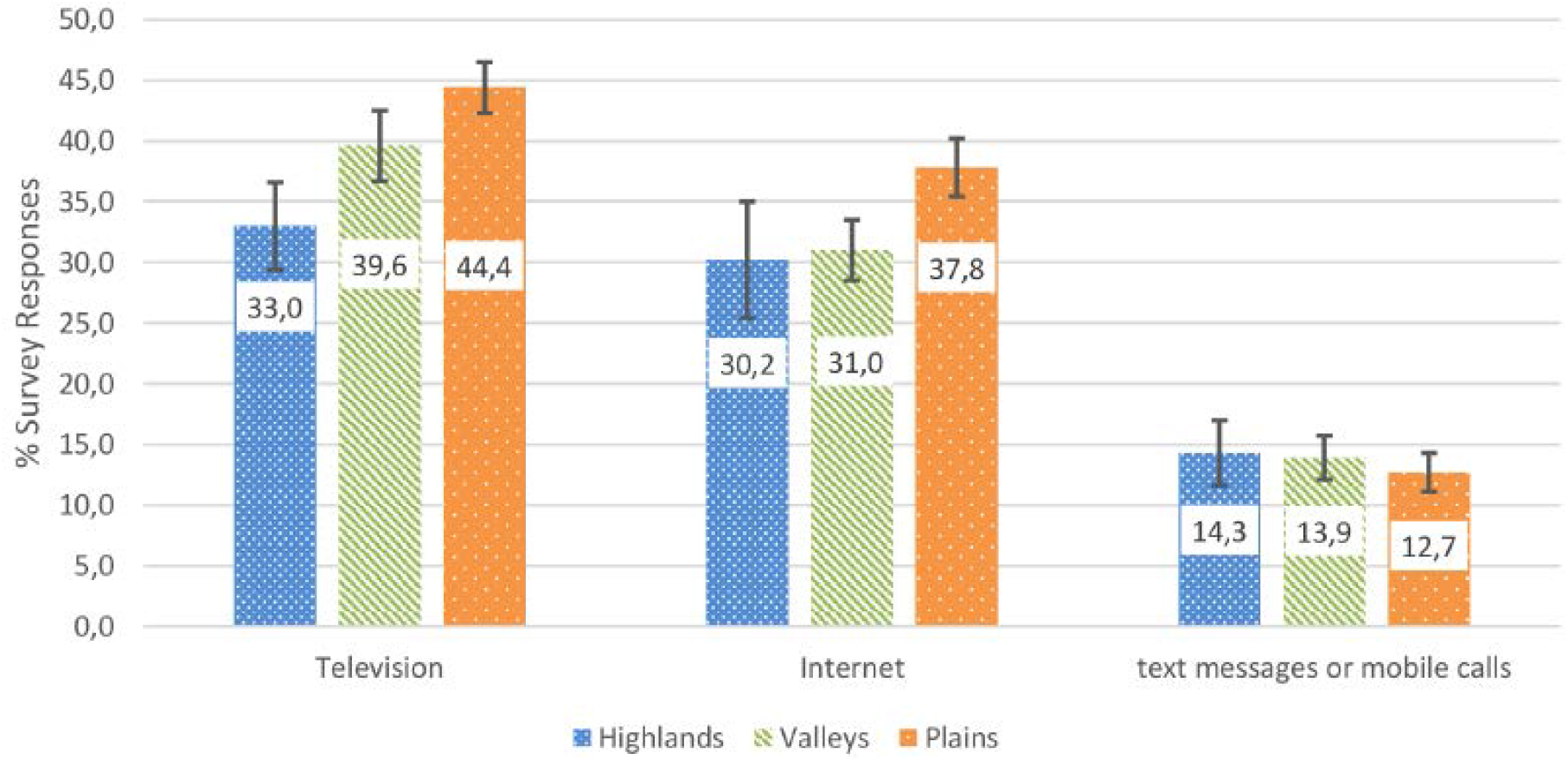
Bolivian students who saw advertisements on junk food and carbonated soft drinks most of the time or always by region

## DISCUSION

Although many countries have applied the GSHS survey, as noted above, they have adapted the questions to their specific culture. We were, therefore, unable to make many national comparisons. Even within Bolivia the GSHS 2012 survey did not include some of questions included in the GSHS 2018 survey.

Breakfast skipping is widely linked to the risk of developing NCD’s such as obesity, diabetes mellitus type 2, and heart diseases (14–18). Although Bolivia has a low percentage of adolescents that never eat breakfast (6,1%), there is also a lower percentage of students who always eat breakfast (53,2%) compared to other countries (Michoacán-Mexico 96%; Cajamarca-Peru: 66,2%) (19,20). Nevertheless, this survey did not explore the quality of the breakfast adolescents consume. Further studies are necessary to determine this.

Patterns on vegetable and fruit consumption show that Bolivian adolescents consume more fruits than vegetables. Similar results were reported in other countries: GSHS Perú 2010: Vegetables 8,9%. Fruits: 31,7%; Ibagué-Colombia: Vegetables 9% Fruits 41% (21,22). The survey we conducted in Bolivia showed that although the highlands has greater poverty and a lower rate of literacy than the valleys and plains, it has a greater presence of protective factors than the other regions. The valleys, despite having greater access to fruits and vegetables because of its climatic characteristics, has a greater presence of risky behaviors. And a comparison between the 2012 and 2018 Bolivia surveys show a reduction in the consumption of vegetables by 2,6% and fruits by19,6% (9). It is urgent to decipher the reason for this decrease so that adequate interventions can be developed.

Junk food and carbonated soft drinks are also highly correlated with increased risk of developing NCDs (23–26). Despite the fact that some departments in Bolivia have already applied politics to limit advertising and diminish the access to junk food and carbonated soft drinks in schools, there is still a high percentage of adolescents who are able to get them at school. Teenagers in the plains have greater exposure to advertising and access to junk food in their schools, which seems to be related to the bad nutritional habits acquired in this region. More effort is needed to comprehend the way adolescents select their food. Some international studies have shown that adolescents are very sensitive to food advertising through the mass media, especially television, which affect their choice, purchase and consumption (27,28).

High access to processed foods and carbonated soft drinks, associated with low consumption of fruits and vegetables are causing overweight, obesity and at the same time micronutrient deficiency. This leads to the so called “double burden” of malnutrition, undernutrition coexisting with overweight and obesity in the same individual (29–31). Undernutrition health effects are linked to impaired childhood development and higher risk of infectious diseases. On the other hand, overweight carries an increased risk of non-communicable diseases. Both lead to higher medical expenses and loss of productivity (32). This economic analysis is critical to support double-duty interventions since evidence shows that single-duty interventions are not efficient and some have even raised the risk of poor-quality diets (33).

Given the traditional variety between the three geographical regions in Bolivia, it was surprising to find that the three geographical regions show similar patterns. Given this finding, it is certain that the same government policy will help the three regions in the same way. Some departments will require specific policies to address specific concerns, but public health policy improvements will be effective for the country.

### Permission to reuse and copyright

Permission to reuse figures, tables, and images may be obtained from the corresponding author.

## Supporting information

STROBE cross-sectional checklist

Disclosure statement

## Data Availability

The raw data supporting the conclusions of this article will be made available by the authors, without undue reservation.

## Author Contributions

Lenildo De Moura, the Ministry of Health () and Marisol Mamani The Institute of Health Investigation designed the study with the Pan American Health Organization’s technical support. Lenildo De Moura, Noelia Villalta and Nattalia Arauco oversaw the data collection, Noelia Villalta performed the statistical analysis and wrote the first draft of the manuscript. Joan Sabaté reviewed the manuscript, reorganized the second draft and contributed to the content. Daniel Illanes and Pramil Singh reviewed and approved the submitted version.

## Acknowledgments

The authors would like to acknowledge the university students and health professionals throughout Bolivia who collected the data. We would also like to acknowledge the cooperation of the schools that participated in the survey.

