## Appendix 1 for "Dietary Patterns Among 13-17-Years Old School Adolescents: A National Comparison of the Three Geographical Regions in Bolivia"

### *2018 BOLIVIA GLOBAL SCHOOL-BASED STUDENT HEALTH SURVEY*

This survey is about your health and the things you do that may affect your health. Students like you all over your country are doing this survey. Students in many other countries around the world also are doing this survey. The information you give will be used to develop better health programs for young people like yourself.

DO NOT write your name on this survey or the answer sheet. The answers you give will be kept private. No one will know how you answer. Answer the questions based on what you really know or do. There are no right or wrong answers.

Completing the survey is voluntary. Your grade or mark in this class will not be affected whether or not you answer the questions. If you do not want to answer a question, just leave it blank.

Make sure to read every question. Fill in the circles on your answer sheet that match your answer. Use only the pencil you are given. When you are done, do what the person who is giving you the survey says to do.

Here is an example of how to fill in the circles:

Fill in the circles like this ● Not like this 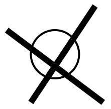 or 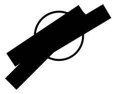

Survey

1. Do fish live in water?
   A. Yes
   B. No

Answer sheet

1. ● Ⓑ Ⓒ Ⓓ Ⓔ Ⓕ Ⓖ Ⓗ

Thank you very much for your help.

1. How old are you?

   A. 11 years old or younger
   B. 12 years old
   C. 13 years old
   D. 14 years old
   E. 15 years old
   F. 16 years old
   G. 17 years old
   H. 18 years old or older

2. What is your sex?

   A. Male
   B. Female

3. In what grade/class/ standard are you?

   A. Second grade of Secondary
   B. Third grade of Secondary
   C. Fourth grade of Secondary
   D. Fifth grade of Secondary
   E. Sixth grade of Secondary

**The next 3 questions ask about your height, weight, and going hungry.**

4. How tall are you without your shoes on?
   ON THE ANSWER SHEET, WRITE YOUR HEIGHT IN THE SHADED BOXES AT THE TOP OF THE GRID. THEN FILL IN THE OVAL BELOW EACH NUMBER.

**Example**

| Height (cm) | | |
|---|---|---|
| ***1*** | ***5*** | ***3*** |
| 0 | 0 | 0 |
| ● | 1 | 1 |
| 2 | 2 | 2 |
| | 3 | ● |
| | 4 | 4 |
| | ● | 5 |
| | 6 | 6 |
| | 7 | 7 |
| | 8 | 8 |
| | 9 | 9 |
| 9 | I do not know | |

5. How much do you weigh without your shoes on? ON THE ANSWER SHEET, WRITE YOUR WEIGHT IN THE SHADED BOXES AT THE TOP OF THE GRID. THEN FILL IN THE OVAL BELOW EACH NUMBER.

**Example**

| Weight (kg) | | |
|---|---|---|
| ***0*** | ***5*** | ***2*** |
| ● | 0 | 0 |
| 1 | 1 | 1 |
| 2 | 2 | ● |
| | 3 | 3 |
| | 4 | 4 |
| | ● | 5 |
| | 6 | 6 |
| | 7 | 7 |
| | 8 | 8 |
| | 9 | 9 |
| 9 | I do not know | |

6. During the past 30 days, how often did you go hungry because there was not enough food in your home?

   A. Never
   B. Rarely
   C. Sometimes
   D. Most of the time
   E. Always

**The next 17 questions ask about what you might eat and drink.**

7. During the past 7 days, how many times did you eat fruit, such as oranges, tangerines, bananas, strawberries, papaya, pineapple, melon, apple, pear, grapes, kiwi, mango, or lime?

   A. I did not eat fruit during the past 7 days
   B. 1 to 3 times during the past 7 days
   C. 4 to 6 times during the past 7 days
   D. 1 time per day
   E. 2 times per day
   F. 3 times per day
   G. 4 or more times per day

8. During the past 7 days, how many times did you eat vegetables, such as lettuce, radishes, carrots, cucumbers, spinach, chard, squash, onions, cauliflower, or broccoli?

   A. I did not eat vegetables during the past 7 days
   B. 1 to 3 times during the past 7 days
   C. 4 to 6 times during the past 7 days
   D. 1 time per day
   E. 2 times per day
   F. 3 times per day
   G. 4 or more times per day

9. During the past 7 days, how many times did you drink a can, bottle, or glass of a carbonated soft drink, such as Pepsi, Coca-Cola, Fanta, Papaya Salvieti or Kinoto? (Do **not** include diet soft drinks.)

   A. I did not drink carbonated soft drinks during the past 7 days
   B. 1 to 3 times during the past 7 days
   C. 4 to 6 times during the past 7 days
   D. 1 time per day
   E. 2 times per day
   F. 3 times per day
   G. 4 or more times per day

10. During the past 7 days, on how many days did you eat food from a fast food restaurant, such as Burger King, Pollos Copacabana, Dumbo, Toby's or Pizza Elli's?

    A. 0 days
    B. 1 day
    C. 2 days
    D. 3 days
    E. 4 days
    F. 5 days
    G. 6 days
    H. 7 days

11. During the past 7 days, how many times did you drink milk or eat milk products, such as yogurt or yokult?

    A. I did not drink milk or eat milk products during the past 7 days
    B. 1 to 3 times during the past 7 days
    C. 4 to 6 times during the past 7 days
    D. 1 time per day
    E. 2 times per day
    F. 3 times per day
    G. 4 or more times per day

12. During the past 7 days, how many times did you eat salty foods, such as salted peanuts?

    A. I did not eat salty foods during the past 7 days
    B. 1 to 3 times during the past 7 days
    C. 4 to 6 times during the past 7 days
    D. 1 time per day
    E. 2 times per day
    F. 3 times per day
    G. 4 or more times per day

13. During the past 7 days, how many times did you eat foods high in fat, such as burgers, fried chicken, or salchipapas?

    A. I did not eat foods high in fat during the past 7 days
    B. 1 to 3 times during the past 7 days
    C. 4 to 6 times during the past 7 days
    D. 1 time per day
    E. 2 times per day
    F. 3 times per day
    G. 4 or more times per day

14. During the past 7 days, how many times did you drink 100% fruit juices, such as orange juice or papaya juice?

    A. I did not drink 100% fruit juices during the past 7 days
    B. 1 to 3 times during the past 7 days
    C. 4 to 6 times during the past 7 days
    D. 1 time per day
    E. 2 times per day
    F. 3 times per day
    G. 4 or more times per day

15. During the past 7 days, how many times did you drink a can, bottle, or glass of a sugar-sweetened beverage such as a sports drink (such as Gatorade), energy drink (such as Red Bull), or fruit drink that was not 100% juice? (Do **not** count carbonated soft drinks or diet drinks)

    A. I did not drink sugar-sweetened beverages during the past 7 days
    B. 1 to 3 times during the past 7 days
    C. 4 to 6 times during the past 7 days
    D. 1 time per day
    E. 2 times per day
    F. 3 times per day
    G. 4 or more times per day

16. During the past 7 days, how many times did you drink **a glass of plain water**, boiled or bottled?

    A. I did not drink water during the past 7 days
    B. 1 to 3 times during the past 7 days
    C. 4 to 6 times during the past 7 days
    D. 1 time per day
    E. 2 times per day
    F. 3 times per day
    G. 4 or more times per day

17. What is your favorite type of drink when you are thirsty?

    A. Water
    B. Sodas, such as Coca Cola, Sprite, Pepsi, or Fanta
    C. Refrescos, such as Mocochinchi or Somó,
    D. Processed juices, such as Frut-all or Nectar
    E. Energy drinks, such as Red Bull
    F. Sports drinks, such as Gatorade or Powerade
    G. Some other type of drink

18. How often do you add sauces, such as mayonnaise, ketchup, or mustard to foods before you try them or while you are eating them?

    A. Never
    B. Rarely
    C. Sometimes
    D. Most of the time
    E. Always

**The next 5 questions ask about eating breakfast and lunch.**

19. During the past 30 days, how often did you eat breakfast?

    A. Never
    B. Rarely
    C. Sometimes
    D. Most of the time
    E. Always

20. During the past 30 days, how often was breakfast offered to you at school?

    A. Never
    B. Rarely
    C. Sometimes
    D. Most of the time
    E. Always

21. What is the **main** reason you do not eat breakfast?

    A. I always eat breakfast
    B. I do not have time for breakfast
    C. I cannot eat early in the morning
    D. There is not always food in my home
    E. Some other reason

22. During the past 30 days, how often did you bring your lunch to school?

    A. Never
    B. Rarely
    C. Sometimes
    D. Most of the time
    E. Always

23. During the past 30 days, how often was lunch offered to you at school?

    A. Never
    B. Rarely
    C. Sometimes
    D. Most of the time
    E. Always

**The next 10 questions ask about your weight and losing or gaining weight.**

24. During the past 12 months, have you been weighed and measured?

    A. Yes
    B. No

25. Which of the following are you trying to do about your weight?

    A. I am **not trying to do anything** about my weight
    B. **Lose** weight
    C. **Gain** weight
    D. **Stay** the same weight

26. During the past 30 days, did you **exercise** to lose weight or to keep from gaining weight?

    A. Yes
    B. No

27. During the past 30 days, did you **take any diet pills, powders, or liquids without a doctor's advice** to lose weight or to keep from gaining weight?

    A. Yes
    B. No

28. During the past 30 days, did you **eat less food, fewer calories, or foods low in fat** to lose weight or to keep from gaining weight?

    A. Yes
    B. No

29. During the past 30 days, did you **go without eating for 24 hours or more** (also called fasting) to lose weight or to keep from gaining weight?

    A. Yes
    B. No

30. During the past 30 days, did you **vomit or take laxatives** to lose weight or to keep from gaining weight?

    A. Yes
    B. No

31. During the past 30 days, did you **exercise** to gain weight?

    A. Yes
    B. No

32. During the past 30 days, did you **eat more food, more calories, or foods high in fat** to gain weight?

    A. Yes
    B. No

33. During the past 30 days, did you **take any pills, powders, or liquids** without a doctor's advice to gain weight?

    A. Yes
    B. No

**The next 7 questions ask about how carbonated soft drinks and foods from fast food restaurants are advertised and sold**.

34. When you watch television, videos, or movies, how often do you see advertisements for carbonated soft drinks or fast foods?

    A. I do not watch television, videos, or movies
    B. Never
    C. Rarely
    D. Sometimes
    E. Most of the time
    F. Always

35. During the past 30 days, how many advertisements for carbonated soft drinks or fast foods did you see when you watched **television**?

    A. I did not watch television during the past 30 days
    B. A lot
    C. A few
    D. None

36. During the past 30 days, how many advertisements for carbonated soft drinks or fast foods did you see on the **internet**?

    A. I did not use the internet during the past 30 days
    B. A lot
    C. A few
    D. None

37. During the past 30 days, how many **text messages or mobile phone calls** did you get that encouraged you to go to a carbonated soft drink or fast food company website?

    A. I did not get text messages or mobile phone calls during the past 30 days
    B. A lot
    C. A few
    D. None

38. During the past 30 days, how many advertisements for carbonated soft drinks or fast foods did you see in your school?

    A. A lot
    B. A few
    C. None

39. Can you buy **carbonated soft drinks** or get them for free in your school?

    A. Yes
    B. No

40. Can you buy **fast foods** or get them for free in your school?

    A. Yes
    B. No

**The next 5 questions ask about what you have learned.**

41. During this school year, were you taught in any of your classes of the benefits of healthy eating?

    A. Yes
    B. No
    C. I do not know

42. During this school year, were you taught in any of your classes of the benefits of eating more fruits and vegetables?

    A. Yes
    B. No
    C. I do not know

43. During this school year, were you taught in any of your classes how to safely prepare or store food?

    A. Yes
    B. No
    C. I do not know

44. During this school year, were you taught in any of your classes healthy ways to gain weight?

    A. Yes
    B. No
    C. I do not know

45. Do you usually read the labels of the food you eat?

    A. Yes
    B. No
