## Supplementary material for "Dietary Patterns Among 13-17-Years Old School Adolescents: A National Comparison of the Three Geographical Regions in Bolivia": Disclosure statement

### ICMJE DISCLOSURE FORM

**Date:**_June 22, 2021_______________________________________________________________________
**Your Name:**_Noelia Villalta______________________________________________________________________
**Manuscript Title:** Dietary Patterns Among 13-17-Years Old School Adolescents: A National Comparison of the Three Geographical Regions in Bolivia
**Manuscript number (if known):**_________________________________________________________________

**In the interest of transparency, we ask you to disclose all relationships/activities/interests listed below that are related to the content of your manuscript. "Related" means any relation with for-profit or not-for-profit third parties whose interests may be affected by the content of the manuscript. Disclosure represents a commitment to transparency and does not necessarily indicate a bias. If you are in doubt about whether to list a relationship/activity/interest, it is preferable that you do so.**

**The following questions apply to the author's relationships/activities/interests as they relate to the current manuscript only.**

**The author's relationships/activities/interests should be defined broadly. For example, if your manuscript pertains to the epidemiology of hypertension, you should declare all relationships with manufacturers of antihypertensive medication, even if that medication is not mentioned in the manuscript.**

**In item #1 below, report all support for the work reported in this manuscript without time limit. For all other items, the time frame for disclosure is the past 36 months.**

| | | **Name all entities with whom you have this relationship or indicate none (add rows as needed)** | **Specifications/Comments (e.g., if payments were made to you or to your institution)** |
|---|---|---|---|
| | | **Time frame: Since the initial planning of the work** | |
| 1 | All support for the present manuscript (e.g., funding, provision of study materials, medical writing, article processing charges, etc.) **No time limit for this item.** | ____ None<br>PAHO/WHO | |
| | | PAHO BOLIVIA | |
| | | CDC | |
| | | IINSAD | |
| | | **Time frame: past 36 months** | |
| 2 | Grants or contracts from any entity (if not indicated in item #1 above). | __X__ None | |
| 3 | Royalties or licenses | __X__ None | |

| | | |
|---|---|---|
| 4 | Consulting fees | __X__ None |
| 5 | Payment or honoraria for lectures, presentations, speakers bureaus, manuscript writing or educational events | __X__ None |
| 6 | Payment for expert testimony | __X__ None |
| 7 | Support for attending meetings and/or travel | __X__ None |
| 8 | Patents planned, issued or pending | __X__ None |
| 9 | Participation on a Data Safety Monitoring Board or Advisory Board | __X__ None |
| 10 | Leadership or fiduciary role in other board, society, committee or advocacy group, paid or unpaid | __X__ None |
| 11 | Stock or stock options | __X__ None |
| 12 | Receipt of equipment, materials, drugs, medical writing, gifts or other services | __X__ None |
| 13 | Other financial or non-financial interests | __X__ None |

**Please place an "X" next to the following statement to indicate your agreement:**

**_X_ I certify that I have answered every question and have not altered the wording of any of the questions on this form.**
